# Social inequalities in youth mental health in Canada, 2007-2022: a population-based repeated cross-sectional study

**DOI:** 10.1101/2024.05.03.24306830

**Authors:** Britt McKinnon, Rabina Jahan, Julia Mazza

## Abstract

**Background:** Rising concern surrounds youth mental health in Canada, with growing disparities between females and males. However, less is known about recent trends by other sociodemographic factors, including sexual orientation, ethnocultural background, and socioeconomic status.

**Methods:** This study analyzed data from 96 683 youths aged 15-24 who participated in the nationally representative Canadian Community Health Survey (CCHS) between 2007 and 2022. Trends in absolute and relative inequalities in poor/fair self-rated mental health (SRMH) by sex, sexual orientation, racialized and Indigenous identity, and socioeconomic conditions were assessed.

**Results:** The percent of youths reporting poor/fair SRMH quadrupled from 4.3% in 2007-08 to 20.1% in 2021-22. During the same period, absolute inequalities in SRMH increased by 9.9 percentage points (95% CI: 6.6, 12.9) for females compared to males, 11.4 percentage points (95% CI: 4.6, 18.2) for Indigenous versus non-racialized youth, and 15.4 percentage points (95% CI: 5.7, 25.1) for youth (aged 18-24) identifying as lesbian, gay, or bisexual (LGB) compared to heterosexual.

**Conclusion:** The sustained deterioration in youth SRMH over the past decade and a half has been accompanied by widening inequalities across several dimensions important for health equity in Canada. Action is needed to identify and implement effective programs and policies to support youth mental health and address disparities.

**What is already known on this topic:**

- Youth mental health has been declining in many countries, including Canada, especially among females
- Trends by other sociodemographic factors, including sexual orientation, ethnocultural background, and socioeconomic status, are less clear

**What this study adds:**

- To our knowledge, this is the first study to examine long-term trends in youth mental health inequalities in Canada over a period marked by significant global events including the 2008 financial crisis and the COVID-19 pandemic.
- Findings show disproportionate declines in mental health among certain youth populations, including those from structurally marginalized backgrounds such as those identifying as lesbian, gay or bisexual (LGB) and Indigenous.

**How this study might affect research, practice or policy:**

- These findings should encourage further research and action to identify and implement evidence-based programs and policies to support youth mental health and reduce disparities.

## Introduction

There is growing global concern about rising mental health problems among young people [1–3]. In Canada, this is evident in the tripling of rates of generalized anxiety disorder and the doubling of rates of major depression among youth aged 15-24 over the past decade [4]. Proposed explanations for the increase include the widespread influence of social media, economic and social stressors, reduced stigma surrounding mental health, and the effects of the COVID-19 pandemic [1,5]. Despite ongoing research efforts, the full scope of factors driving these trends remains incompletely understood, yet the urgency of addressing this issue is undeniable, given the potential long-term and life course consequences of early mental health challenges [2,3].

Social determinants, including economic deprivation, family and peer relationships, discrimination and social exclusion, and exposure to stressful environments, have a profound influence on youth mental health [2,3]. In Canada, mental health challenges and disorders have been shown to disproportionately affect females, LGBTQ+ youth, Indigenous youth, and those from socioeconomically disadvantaged backgrounds [3,4,6,7]. There is evidence that youth mental health has been deteriorating more rapidly among females than males [4], yet less is known about recent inequality trends by other sociodemographic factors.

To address this gap, this study examined trends in mental health inequalities among youth in Canada between 2007 and 2022, a period marked by significant global events including the 2008 financial crisis and the COVID-19 pandemic. We assess absolute and relative inequalities based on characteristics meaningful to health equity in Canada, including sex/gender, socioeconomic conditions, racialized and Indigenous identity, and sexual orientation.

## Methods

### Study population

The study analyzed data from 96 683 individuals aged 15-24 who participated in the Canadian Community Health Survey (CCHS) between 2007 and 2022. The CCHS is an annual nationally representative, cross-sectional survey of the Canadian population aged 12 and older, excluding those living on reserves, active-duty military, and institutionalized persons (approximately 3% of the Canadian population). Survey response rates for the years examined ranged from 49% to 70%, except for substantially lower rates during the COVID-19 pandemic (29% in 2020 and 24% in 2021). Additionally, CCHS underwent redesigns of its sampling methodology and content in both 2015 and 2022. Statistics Canada provides sampling and bootstrap replication weights to account for unequal sampling selection probabilities and non-response.

### Measures

The primary outcome of self-rated mental health (SRMH) was measured by the question: “In general would you say your mental health is: excellent? very good? good? fair? poor?” Responses were categorized as fair/poor and good/very good/excellent. Unlike several other mental health outcomes (e.g., psychological distress, depressive symptoms), SRMH has been included consistently across CCHS waves and is a key outcome in Canadian public health surveillance of mental health, including among youth [8]. Canadian evidence demonstrates that SRMH is strongly associated with psychological distress and major depression among the general population [9] and with depressive symptoms among adolescents [10]. A longitudinal study of Canadian youth also identified SRMH as a significant predictor of depressive symptoms in midlife [11].

Respondents reported their age in years (categorized as 15-19, 20-24); sex (male, female); sexual orientation (categorized as heterosexual; lesbian, gay or bisexual (LGB); and visible minority [12] and Indigenous identity [13] as defined by Statistics Canada (categorized as non-racialized; non-Indigenous racialized; Indigenous). The area-based socioeconomic measure was based on 2006 and 2016 versions of the Canadian Marginalization Index of neighborhood material resources, which incorporates indicators representing access to and attainment of basic material needs (e.g., % of population unemployed, % of population below low-income cut-off) [14]. This indicator was preferred to household income because the 15-24 demographic often experiences a transition from living in a parent’s household to living independently.

### Statistical analysis

We estimated the prevalence of poor/fair SRMH by sociodemographic characteristics from logistic regression models followed by the Stata margins postestimation command [15]. Interaction terms between sociodemographic variables and time (2-year intervals) assessed whether changes in the prevalence of low SRMH differed by sex, LGB identity, racialized identity, neighbourhood deprivation, and age groups. Absolute inequalities in SRMH were measured by prevalence differences (PD) and relative inequalities by prevalence ratios (PR). Point estimates and 95% confidence intervals incorporated CCHS sampling and bootstrap replication weights.

## Results

The prevalence of poor/fair SRMH increased by 15.8 percentage points (95% confidence interval (CI): 14.3, 17.4) from 4.3% in 2007-08 to 20.1% in 2021-22 (Figure 1, panel A, presents annual trends in poor/fair SRMH). Two-year prevalence trends stratified by sociodemographic groups are presented in Figure 1, panels B to F, and corresponding estimates for 2007-08 and 2021-22 are shown in Table 1. Significant increases in fair/poor SRMH were evident across all sociodemographic groups, ranging from a 11.1 percentage point rise among male youth (95% CI: 9.0, 13.1) to a 28.1 percentage point rise among youth identifying as LGB (95% CI: 21.2, 34.4). In 2021-22 compared to 2007-08, the sample was substantially more likely to identify as LGB (12.7% vs. 2.5%) and non-Indigenous racialized (31.6% vs 16.0%) (Table 1).

**Table 1:** Trends in estimated prevalence, prevalence differences (PD), and prevalence ratios (PR) for poor/fair SRMH by sociodemographic characteristics, Canadian youth aged 15-24 years

|  | 2007-08 (N = 14 984) |  |  |  | 2021-22 (N = 7403) |  |  |  | Change 2007-08 to 2021-22 |  |  |
| --- | --- | --- | --- | --- | --- | --- | --- | --- | --- | --- | --- |
|  | % of sample | Prevalence (95% CI) | PD (95% CI) | PR (95% CI) | % of sample | Prevalence (95% CI) | PD (95% CI) | PR (95% CI) | Prevalence (95% CI) | PD (95% CI) | PR (95% CI) |
| Overall sample | - | 4.3 (3.8, 4.9) | - | - | - | 20.2 (18.8, 21.6) | - | - | 15.9 (14.4, 17.4) | - | - |
| Age |  |  |  |  |  |  |  |  |  |  |  |
| 15-19 years | 48.6 | 4.1 (3.5, 4.8) | 0 (ref) | 1 (ref) | 50.9 | 18.2 (16.6, 19.8) | 0 (ref) | 1 (ref) | 14.1 (12.3, 15.8) | - | - |
| 20-24 years | 51.4 | 4.5 (3.6, 5.3) | 0.3 (-0.7, 1.4) | 1.08 (0.82, 1.34) | 49.1 | 22.1 (19.6, 24.6) | 3.9 (0.9, 6.9) | 1.21 (1.04, 1.39) | 17.6 (15.0, 20.3) | 3.6 (0.4, 6.7) | 1.13 (0.81, 1.44) |
| Sex |  |  |  |  |  |  |  |  |  |  |  |
| Male | 51.4 | 3.9 (3.2, 4.7) | 0 (ref) | 1.0 (ref) | 52.0 | 15.0 (13.1, 16.9) | 0 (ref) | 1 (ref) | 11.1 (9.0, 13.1) | - | - |
| Female | 48.6 | 4.7 (3.9, 5.5) | 0.8 (-0.3, 1.8) | 1.20 (0.91, 1.49) | 48.0 | 25.7 (23.7, 27.7) | 10.7 (7.9, 13.5) | 1.71 (1.45, 1.97) | 21.0 (18.8, 23.2) | 9.9 (6.9, 12.9) | 1.43 (1.02, 1.83) |
| Sexual orientation (age 18-24)* |  |  |  |  |  |  |  |  |  |  |  |
| Heterosexual | 97.5 | 4.3 (3.675, 5.0) | 0 (ref) | 1.0 (ref) | 87.3 | 17.0 (15.1, 18.9) | 0 (ref) | 0 (ref) | 12.7 (10.7, 14.7) | - | - |
| Gay/lesbian/bisexual | 2.5 | 13.6 (6.9, 20.3) | 9.3 (2.6, 15.9) | 3.16 (1.57, 4.740) | 12.7 | 41.7 (34.8, 48.6) | 24.7 (17.5, 31.9) | 2.45 (1.95, 2.95) | 28.1 (18.7, 37.5) | 15.4 (5.7, 25.1) | 0.78 (0.36, 1.19) |
| Racialized identity |  |  |  |  |  |  |  |  |  |  |  |
| Non-racialized | 79.0 | 4.1 (3.5, 4.7) | 0 (ref) | 1.0 (ref) | 63.8 | 20.4 (18.6, 22.3) | 0 (ref) | 1.0 (ref) | 16.4 (14.4, 18.3) | - | - |
| Non-Indigenous racialized | 16.0 | 5.2 (3.3, 7.1) | 1.1 (-0.9, 3.1) | 1.27 (0.77, 1.76) | 31.6 | 17.5 (14.8, 20.3) | -2.9 (-6.3, 0.5) | 0.86 (0.70, 1.02) | 12.4 (9.1, 15.6) | -4.0 (-7.8, -0.2) | 0.68 (0.39, 0.96) |
| Indigenous | 5.0 | 5.5 (3.6, 7.3) | 1.4 (-0.6, 3.3) | 1.34 (0.84, 1.83) | 4.6 | 33.3 (27.0, 39.6) | 12.8 (6.4, 19.3) | 1.63 (1.29, 1.96) | 27.8 (21.2, 34.4) | 11.4 (4.6, 18.2) | 1.29 (0.72, 1.85) |
| Neighbourhood socioeconomic conditions |  |  |  |  |  |  |  |  |  |  |  |
| Less deprived (top 60%) | 63.2 | 4.4 (3.7, 5.0) | 0 (ref) | 1 (ref) | 68.4 | 20.6 (18.9, 22.4) | 0 (ref) | 1.0 (ref) | 16.3 (14.4, 18.2) | - | - |
| More deprived (lower 40%) | 36.8 | 4.2 (3.4, 5.0) | -0.2 (-1.2, 0.9) | 0.97 (0.74, 1.20) | 31.6 | 19.1 (16.6, 21.6) | -1.6 (-4.7, 1.6) | 0.92 (0.78, 1.07) | 14.9 (12.2, 17.5) | -1.4 (-4.7, 1.9) | 0.96 (0.69, 1.23) |

Note: Estimates are from average marginal effects calculated after logistic regression with bootstrap standard errors. Sexual orientation analysis applies to youth aged 18-24 (N=58 750). Indigenous and racialized identity were self-reported. The “racialized, non-Indigenous” group is equivalent to Statistics Canada’s definition of visible minority as “persons, other than Aboriginal peoples, who are non-Caucasian in race or non-white in colour” [12].

**Figure 1:**
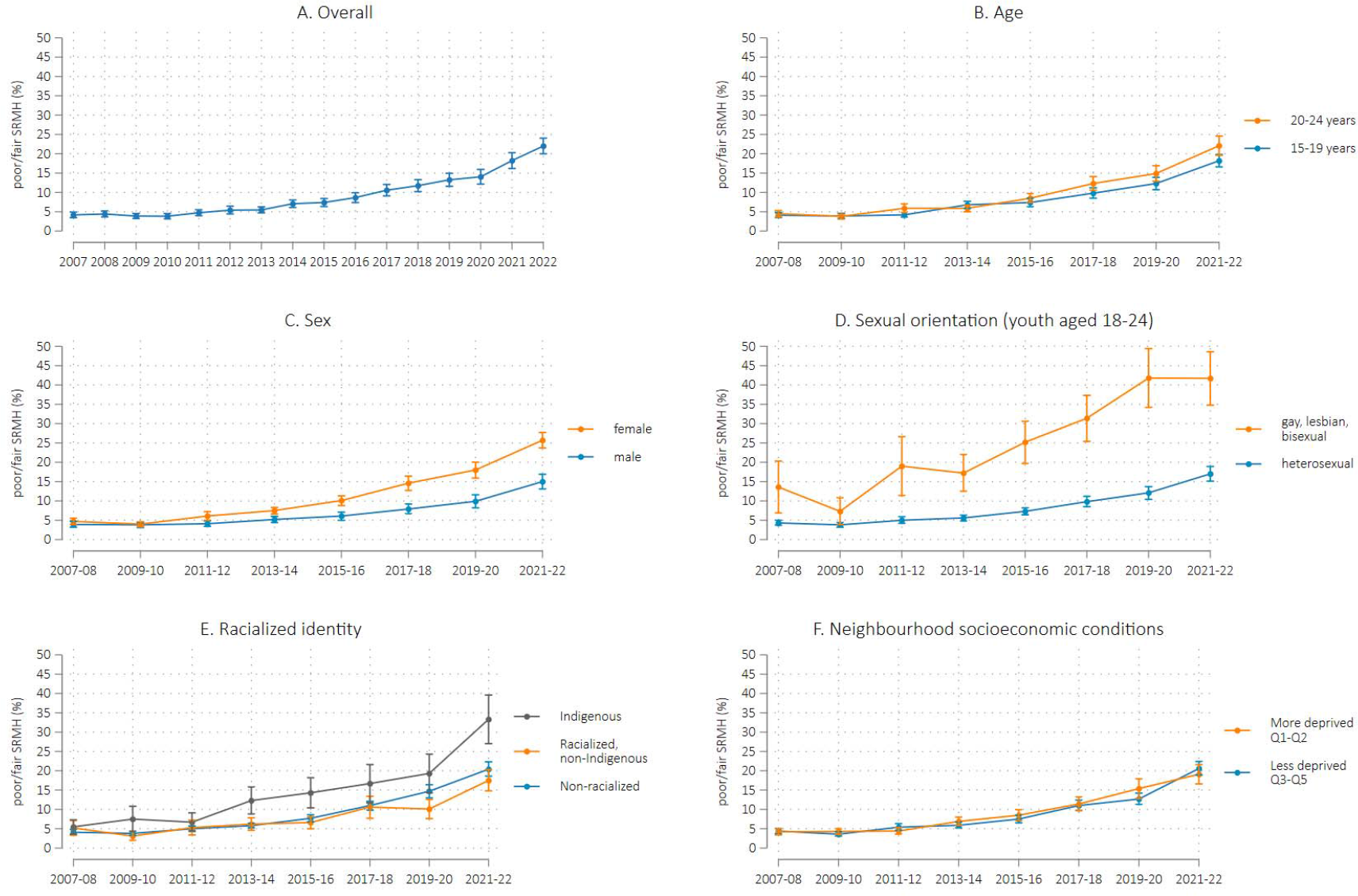
Prevalence trends for poor/fair self-rated mental health by sociodemographic characteristics, Canadian youth aged 15-24 years Note: Estimates are from average marginal effects calculated after logistic regression with bootstrap standard errors. Sexual orientation analysis applies to youth aged 18-24 (N=58 750). Indigenous and racialized identity were self-reported. The “racialized, non-Indigenous” group is equivalent to Statistics Canada’s definition of visible minority as “persons, other than Aboriginal peoples, who are non-Caucasian in race or non-white in colour” [12].

Prevalence differences show widening absolute inequality in fair/poor SRMH by sex, age group, Indigenous identity, and sexual orientation among young people (Table 1). Between 2007-08 and 2021-22, the gap in fair/poor SRMH increased by 9.9 percentage points (95% CI: 6.9, 12.9) for females vs. males, 3.6 percentage points (95% CI: 0.4, 6.7) for youth aged 20-24 vs. 15-19, 11.4 percentage points (95% CI: 4.6, 18.2) for Indigenous vs. non-racialized youth, and 15.4 percentage points (95% CI: 5.7, 25.1) for youth identifying as LGB vs. heterosexual. Neighbourhood-level socioeconomic status showed little association with youth SRMH. The only declining absolute inequality trend was between non-Indigenous racialized and non-racialized youth (PD change of -4.0 percentage points; 95% CI: -7.8, -0.2).

Relative inequalities in fair/poor SRMH were also evident across several sociodemographic factors, especially in 2021-22. However, in general, relative inequality trends did not parallel absolute inequality trends, reflecting the substantial increase in fair/poor SRMH across all groups. PR estimates were consistent with increasing relative inequality trends by sex, age group, and Indigenous identity, although some 95% Cis included the null value. Relative inequality between non-Indigenous racialized youth and non-racialized youth declined by 32% over time (95% CI: 4%, 61%), which was consistent with the declining absolute inequality trend.

## Discussion

In this nationally representative study of over 90 000 youth aged 15-24 years, the prevalence of poor/fair SRMH more than quadrupled from 4.3% to 20.1% between 2007-08 and 2021-22. This trajectory of deteriorating youth mental health seemed to increase in 2020-22, but was well underway prior to the onset of the COVID-19 pandemic.

Our results confirmed a widening gap in SRMH between females and males, driven largely by the more than 5-fold increase in fair/poor SRMH among females over the examined period. This is consistent with evidence of a growing disparity in depression, anxiety, and internalizing symptoms between females and males in several high-income countries, including Canada [1,4]. Plausible explanations for the disproportionate mental health declines among females include the influence of digital media, cyber-bullying, academic pressures, and earlier emergence of puberty among females [1,5].

We also found evidence of large and widening SRMH inequality by sexual orientation among young people, with the gap between LGB and heterosexual youth more than doubling from 8.9 to 22.5 percentage points over the decade and a half. The increased risk of mental ill-health among sexual minority youths is well-documented, attributed primarily to heightened stress stemming from discrimination, bullying, lack of social support, and identity struggles [16,17]. Further investigation of this widening mental health gap between LGB and heterosexual youths is warranted, particularly considering Canada’s comprehensive legal protections for sexual and gender minorities [18].

The enduring intergenerational impacts of colonization impose unique structural inequities on Indigenous youth, contributing significantly to mental health disparities [7]. The widening gap in mental health between Indigenous and non-racialized youth underscores the necessity of investing in mental health interventions that “transcend the Western biomedical model, use a strengths-based approach, and account for the cultural practices and belief systems of Indigenous people” [19].

The current study is limited in its reliance on SRMH, a subjective measure lacking specificity as a measure of psychological distress or mental disorder. Nonetheless, SRMH remains widely endorsed for monitoring population mental health [8,20] and supplementary analyses aligning youth SRMH trends with self-reported mood and anxiety disorder diagnoses bolster its validity (see Figure S1). Limitations in sociodemographic measures and sample size considerations hindered a comprehensive examination of mental health trends among gender diverse or non-binary youths, specific racialized or Indigenous populations, and the intersections of different social identities. Finally, utilizing repeated cross-sectional data with fluctuations in response rates and methodology over the 16-year period raises the possibility that trends may reflect differences in surveyed populations over time, although this likelihood is mitigated by incorporating survey weights.

Findings from this study suggest a need for further research to examine the factors contributing to the disproportionate declines in mental health among certain youth populations, especially those from structurally marginalized backgrounds like those identifying as LGB or Indigenous. Additionally, action is needed to both identify and implement evidence-based programs and policies aimed at addressing mental health disparities among youths.

## Data Availability

All data produced in the present work are contained in the manuscript

## References

1 Keyes KM, Platt JM. Annual Research Review: Sex, gender, and internalizing conditions among adolescents in the 21st century – trends, causes, consequences. J Child Psychol Psychiatry. Published Online First: 2023. doi: 10.1111/jcpp.13864

2 Thapar A, Eyre O, Patel V, et al. Depression in young people. Lancet. 2022;400:617–31.

3 Malla A, Shah J, Iyer S, et al. Youth Mental Health Should Be a Top Priority for Health Care in Canada. Can J Psychiatry. 2018;63:216–22.

4 Stephenson E. Mental disorders and access to mental health care. Statistics Canada 2023. https://www150.statcan.gc.ca/n1/pub/75-006-x/2023001/article/00011-eng.pdf (accessed 11 October 2023)

5 Twenge JM, Martin GN. Gender differences in associations between digital media use and psychological well-being: Evidence from three large datasets. J Adolesc. 2020;79:91–102.

6 Veale JF, Watson RJ, Peter T, et al. Mental Health Disparities Among Canadian Transgender Youth. J Adolesc Heal. 2017;60:44–9.

7 Anderson T. Portrait of youth in Canada: Data report?; Chapter 4: Indigenous Youth in Canada. Statistics Canada 2021. https://www150.statcan.gc.ca/n1/en/pub/42-28-0001/2021001/article/00004-eng.pdf?st=A-Cdhb1L (accessed 26 March 2024)

8 Orpana H, Vachon J, Dykxhoorn J, et al. Monitoring positive mental health and its determinants in Canada: the development of the Positive Mental Health Surveillance Indicator Framework. Heal Promot Chronic Dis Prev Can. 2016;36:1–10.

9 Mawani FN, Gilmour H. Validation of self-rated mental health. Heal Rep. 2010;21:61–75.

10 Sawatzky R, Ratner PA, Johnson JL, et al. Self-reported physical and mental health status and quality of life in adolescents: a latent variable mediation model. Heal Qual Life Outcomes. 2010;8:17.

11 Galambos NL, Johnson MD, Krahn HJ. Self-rated mental health in the transition to adulthood predicts depressive symptoms in midlife. Curr Psychol. 2023;42:30223–34.

12 Statistics Canada. Visible minority of a person. https://www23.statcan.gc.ca/imdb/p3Var.pl?Function=DEC&Id=45152 (accessed 19 March 2024)

13 Statistics Canada. Indigenous identity of person. https://www23.statcan.gc.ca/imdb/p3Var.pl?Function=DEC&Id=42927 (accessed 19 March 2024)

14 Matheson FI, Dunn JR, Smith KLW, et al. Development of the Canadian Marginalization Index: a new tool for the study of inequality. Can J public Heal Rev Can sante publique. 2011;103:S12–6.

15 Muller CJ, MacLehose RF. Estimating predicted probabilities from logistic regression: different methods correspond to different target populations. International Journal of Epidemiology. 2014;43:962–70.

16 Lucassen MF, Stasiak K, Samra R, et al. Sexual minority youth and depressive symptoms or depressive disorder: A systematic review and meta-analysis of population-based studies. Aust N Zealand J Psychiatry. 2017;51:774–87.

17 Amos R, Manalastas EJ, White R, et al. Mental health, social adversity, and health-related outcomes in sexual minority adolescents: a contemporary national cohort study. Lancet Child Adolesc Heal. 2020;4:36–45.

18 OECD. Over the Rainbow? The Road to LGBTI Inclusion. 2020. 10.1787/8d2fd1a8-en. (accessed 29 March 2024)

19 Carrier L, Shin HD, Rothfus MA, et al. Protective and resilience factors to promote mental health among Indigenous youth in Canada: a scoping review protocol. BMJ Open. 2022;12:e049285.

20 OECD. Measuring Population Mental Health. 2023. 10.1787/5171eef8-en. (accessed 2 April 2024)

