## Supplemental Table 1 for "Social inequalities in youth mental health in Canada, 2007-2022: a population-based repeated cross-sectional study"

Supplementary Material

Table S1: Trends in the prevalence of fair/poor self-rated mental health, self-reported mood disorder diagnosis, and self-reported anxiety disorder diagnosis among youth aged 15-24 years, Canadian Community Health Survey 2007-2022

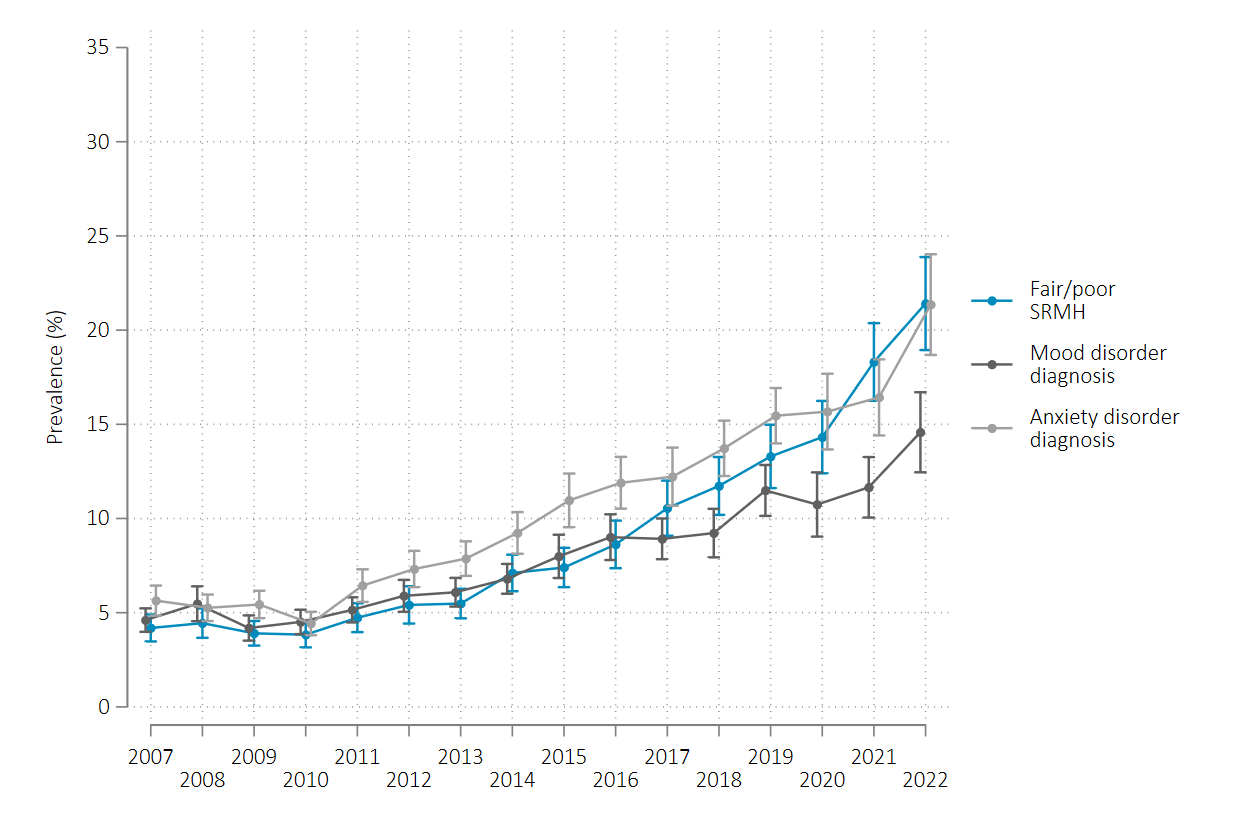

Note: **Self-reported mood disorder diagnosis**: proportion of youth aged 15-24 who reported that they have been diagnosed by a health professional as having a mood disorder, such as depression, bipolar disorder, mania or dysthymia**. Self-reported anxiety disorder diagnosis**: proportion of youth who reported that they have been diagnosed with an anxiety disorder such as a phobia, obsessive-compulsive disorder or a panic disorder.
